# Diagnosis of Pathological Speech with Efficient and Effective Features for Long Short-Term Memory Learning

**DOI:** 10.1101/2023.09.04.23295008

**Authors:** Tuan D. Pham, Simon B. Holmes, Lifong Zou, Mangala Patel, Paul Coulthard

## Abstract

The majority of voice disorders stem from improper vocal usage. Alterations in voice quality can also serve as indicators for a broad spectrum of diseases. Particularly, the significant correlation between voice disorders and dental health underscores the need for precise diagnosis through acoustic data. This paper introduces effective and efficient features for deep learning with speech signals to distinguish between two groups: individuals with healthy voices and those with pathological voice conditions. Using a public voice database, the ten-fold test results obtained from long short-term memory networks trained on the combination of time-frequency and time-space features with a data balance strategy achieved the following metrics: accuracy = 90%, sensitivity = 93%, specificity = 87%, precision = 88%, *F*_1_ score = 0.90, and area under the receiver operating characteristic curve = 0.96.

## 1 Introduction

The involvement of speech pathology in head and neck cancer, and oral-maxillofacial surgery has recently been highlighted by the Michigan Medicine [1]. It is known that voice disorders stem from a wide array of factors, either present since birth or acquired later in life. These factors encompass physiological or anatomical irregularities in the upper airway due to underlying neurological conditions, trauma, and head and neck cancer.

Roughly 30% of adults might encounter difficulties linked to voice disorders [1]. The consequences of voice disorders on quality of life can lead to social seclusion and hinder one’s professional and personal pursuits. The field of speech-language pathology also encompasses the specialized management of head and neck disorders. Within this domain, individualized assessment, treatment, and education are provided to patients dealing with diseases or dysfunctions in the structures of the head and neck, which includes the mandible, maxilla, soft and hard palates, nose, cheek, lips, tongue, and throat [1].

The development of speech within the evolving craniofacial complex relies significantly on the structural and functional integrity of this complex [2]. While development initiates during prenatal stages, a substantial portion of communicative language potential unfolds postnatally and hinges on neurological and cognitive well-being. Both genetic and acquired abnormalities can lead to significant dental misalignments, potentially impacting speech production. Certain investigations indicated a connection between temporomandibular disorders, voice, and oral health-related quality of life in women [3]. Thus, addressing maxillofacial disorders typically necessitates a combined approach involving orthognathic surgery and orthodontic interventions [4]. Optimal patient care requires a truly interdisciplinary approach, ensuring comprehensive and well-coordinated treatment. Effective and efficient rehabilitation plans and schedules can be crafted when speech pathologists and maxillofacial surgeons collaborate closely to synchronize their treatment efforts [2, 5, 6].

In fact, oral health challenges in the field of speech pathology were pointed out decades ago [7]. Assessment of the spectral characteristics of the vowels /a/, /i/, and /u/ was carried out in a group of 24 children [8]. The authors found statistically significant variations in fundamental frequency when analyzing the individual progress of patients with bilateral cleft lip, jaw, and palate conditions. The fundamental frequency and analysis of the first formant demonstrated their effectiveness in characterizing the vocal timbre of individuals with cleft conditions.

In pediatric dentistry, it was investigated that children with voice disorders tend to exhibit a greater prevalence and more pronounced malocclusions than those with ordinary speech development [9]. Moreover, dental medicine is deeply involved in modifying and repairing oral structures to combat the effects of disease and developmental irregularities. Given that a significant part of speech articulation occurs within the oral cavity, any modifications or restorations of these structures can impact speech, the extent of which depends on the location and extent of the alteration. A literature review highlighted the role of pediatric dentists in the early identification and management of speech impairments [10].

The association between speech disorders and dental medicine addressed above highlights the importance of developing a capacity for precise diagnosis of pathological speech. It has been realized that physicians currently face a deficit in diagnostic tools for voice disorders, and the incorporation of artificial intelligence (AI) tools into their toolkit has the potential to expedite diagnoses [11]. A recent review on machine learning methods for diagnosing voice disorders has also been reported with a focus on nonlaryngeal aerodigestive and neurological disorders [12].

Most recently, to tackle this issue, a recent work [13] has suggested the use of a transfer learning framework, which combined a pre-trained convolutional neural network called OpenL3 with a support vector machine classifier for the automatic identification of multiclass voice disorders. Initially, the Mel spectrum of the provided voice signal was extracted, similar to traditional speech feature extraction methods. Subsequently, this Mel spectrum was fed into the OpenL3 network to generate high-level feature embeddings. Powered by deep learning in AI, to distinguish between healthy and pathological voices through the analysis of spectrogram images obtained from the recordings of the vowel /a/, a convolutional neural network model was integrated into a mobile health application, enabling a user-friendly and portable tool for evaluating voice disorders [14]. An alternative approach [15] combined three vocal attributes: chroma, mel spectrogram, and mel frequency cepstral coefficient. The researchers employed a deep neural network to detect voice disorders, utilizing the vowels /a/, /i/, and /u/ articulated at various pitch levels–high, low, and average.

This study presents a substantial extension of a recent work [16] on the selection of efficient and effective features for long short-term memory (LSTM) network learning on speech signals for diagnosing voice disorders presented at the *2023 IEEE Conference on Artificial Intelligence*. Here, the proposed approach explores various feature extraction techniques to address the computational challenges associated with the LSTM. These features encompass derivations from both time-frequency and time-space domains, as well as wavelet time scattering networks.

The rest of this paper is organized as follows. Section 2 outlines the voice dataset used, consisting of both healthy and pathological cases. Section 3 details the techniques applied for extracting features and classifying patterns in the diagnosis of voice disorders. The outcomes of testing multiple models for the diagnosis are presented in Section 4. Finally, Section 5 provides concluding insights from this research, along with potential avenues for future exploration.

## 2 Data

In this study, the VOICED (VOice ICar fEDerico II) database [17] was employed to demonstrate the effectiveness of the proposed approach in addressing a challenging biomedical classification problem. The database comprises acoustic recordings of the vowel “a” lasting five seconds, featuring two distinct groups of participants: healthy individuals and those with pathological conditions. Both groups consist of male and female subjects ranging in age from 18 to 70 years.

Specifically, the dataset contains 61 recordings from healthy voices and 147 from pathological voices, with variable signal lengths about 35,000 time points. Among these, 21 recordings pertain to healthy male voices and 51 to pathological male voices. For female voices, there are 40 healthy voice recordings and 96 pathological voice recordings. To ensure the reliability of the dataset, both healthy and pathological voices underwent rigorous clinical evaluation by medical experts. Diagnoses adhered to the guidelines outlined in the SIFEL protocol, which is a clinical protocol endorsed by the Italian Society of Phoniatrics and Logopaedics.

All recordings took place in a noise-free environment, using Vox4Health technology. The distance between the mobile device and the subjects was approximately 20 cm, at an angle of approximately 45 degrees. All participants received instructions to articulate the vowel in a natural manner, and the acquired signals were filtered to eliminate noise during the recording process. The database includes voice recordings featuring various forms of pathological conditions, which are classified as 1) hyperkinetic dysphonia, 2) hypokinetic dysphonia, and 3) reflux laryngitis. These types of pathology are briefly described as follows [18]:

- *Hyperkinetic dysphonia* represents a frequently encountered clinical condition, especially among individuals engaged in vocally demanding professions. This disorder is marked by excessive muscular contractions within the pneumo-phonic apparatus. It results in a strained, high-pitched voice, diminished frequency modulation, and a noticeable harshness in vocal delivery. Additionally, the increased resistance of the vocal folds to the expiratory airflow intensifies the effort required for phonation, leading to disruptions in respiratory patterns. Numerous conditions fall under this category, including vocal fold nodules, Reinke’s edema, chorditis, rigid vocal folds, polyps, and prolapse.
- *Hypokinetic dysphonia* is characterized by reduced vocal fold adduction during the respiratory cycle, especially during inhalation, resulting in airflow obstruction within the larynx. This incomplete vocal fold closure results in a weak and breathy voice. Interestingly, in cases of hypokinetic dysphonia, voice quality improves with increased vocal intensity, which can potentially lead to improper vocal strain. Conditions falling under the umbrella of hypokinetic dysphonia include dysphonia of the vocal fold groove, adduction deficits, presbiphonia, glottic insufficiency, vocal fold paralysis, conversion dysphonia, laryngitis, and extraglottic air leakage.
- *Reflux laryngitis* refers to an inflammation of the larynx triggered by the regurgitation of stomach acid into the esophagus. The primary symptom typically observed is persistent hoarseness, although additional symptoms may manifest to varying degrees, including pharyngitis, occasional coughing fits, nighttime coughing, asthma, nocturnal laryngeal spasms, and halitosis.

## 3 Methods

The process of extracting time-frequency and time-space features from time series data for LSTM-based classification was initially introduced in [19], and is further elaborated upon here. Additionally, the fundamental concept of wavelet time scattering as a feature extraction technique is introduced to provide insights into the implementation of the proposed approach for diagnosing pathological speech signals.

### 3.1 Extraction of time-frequency features with instantaneous frequency and spectral entropy

The instantaneous frequency (IF) of a non-stationary signal is a time-dependent parameter that corresponds to the mean of the frequencies, denoted as *f*, within the evolving signal as it progresses through various time points *t* [20]. To estimate the IF of a signal at a given sampling rate, the IF function calculates the power spectrum of the spectrogram, denoted as *P*(*t, f*), and then estimates the IF using the following expression:

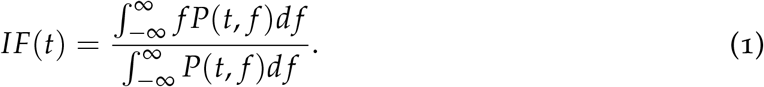

The power spectrum quantifies the strength of a signal at a specific frequency *f*. In the case of a periodic signal, peaks are observed at the fundamental frequency and its harmonics within the spectrum. Quasiperiodic signals exhibit peaks at linear combinations of related frequencies, while chaotic signals result in the presence of broad-band components in the spectrum. However, in practical scenarios, it is impossible to determine the exact power spectrum because real signals are not infinitely long but instead measured over a finite time interval. Consequently, it becomes necessary to estimate the power spectrum numerically and was technically described in [19].

Spectral entropy (SE) of a signal provides insight into its spectral power distribution [20]. The SE treats the normalized power distribution in the frequency domain as a probability distribution and computes its Shannon entropy. In this context, the Shannon entropy is referred to as the spectral entropy of the signal. The probability distribution at a specific time *t*, where 0 ≤ *t* ≤ *T*, and frequency point *z*, denoted as *p*(*t, z*), is computed as follows.

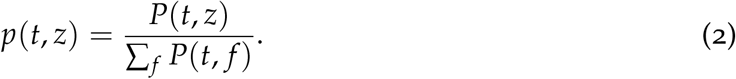

The spectral entropy at time *t*, denoted as *SE*(*t*), is determined as

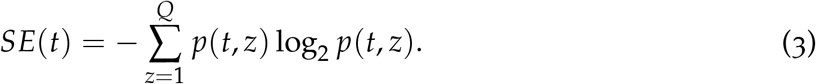

where *Q* is the total frequency points.

In this study, to extract the IF and SE features of the speech signals, the parameters were specified as follows: range of *f* = [0, *f s*/2], and sampling rate *f s* = 300 Hz.

### 3.2 Extraction of time-space features with fuzzy recurrence plots and spatial entropy

A fuzzy recurrence plot (FRP) [21] is a visualization technique used in the analysis of time series data. It is derived from the concept of recurrence plots (RPs) [22], which are used to study nonlinear dynamics in time series. FRPs extend this idea by introducing a degree of fuzziness or uncertainty into the recurrence analysis.

A traditional RP is constructed by comparing each point in the phase space of a dynamical system with every other point and determining whether they are close enough based on some predefined distance metric. If two points are sufficiently close, a black dot that represents “recurrence” is marked on the 2D plot. This is typically represented as a binary plot, where a recurrence is denoted as a black point and non-recurrence as a white point. In an FRP, the binary nature of recurrence is relaxed. Instead of just marking points as either recurrent or non-recurrent, it assigns a degree of membership to each point, representing the degree of similarity or recurrence. This introduces a level of fuzziness or uncertainty into the analysis. The degree of membership takes real values between 0 and 1, where 0 means no recurrence (completely dissimilar) and 1 means a perfect match (completely similar). Real values between 0 and 1 indicate the level of partial similarity. FRPs can be useful for analyzing time series data when the distinction between recurrence and non-recurrence is inherently not clear-cut. This uncertainty modeling allows for a more natural understanding of the data and can reveal hidden patterns or relationships that might not be apparent in traditional binary RPs.

Given an embedding dimension *d* and a time delay *β*, a phase-space construction of the original time series or sequence (*x*_1_, *x*_2_, …, *x*_*M*_) yields **Y** = (**y**_1_, …, **y**_*N*_), where *N* = *M* − (*d* − 1)*β*. The constructed elements of **Y** can be expressed as follows:

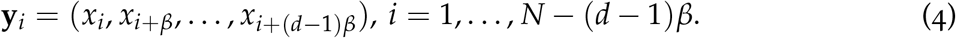

Using this constructed phase space **Y**, an FRP, denoted as **R**, can computed and represented as a grayscale image to visualize the recurrence patterns of a dynamical system. More precisely, an FRP is a square matrix containing membership grades that quantify the similarity between pairs of points in the constructed phase-space trajectory of the dynamical system. This similarity is mathematically expressed as:

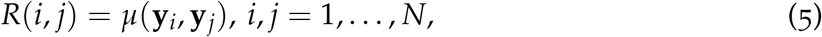

where *μ*(**y**_*i*_, **y**_*j*_) ∈ [0, 1] represents the membership of similarity between **y**_*i*_ and **y**_*j*_. A higher value of *μ*(**y**_*i*_, **y**_*j*_) suggests a stronger similarity between **y**_*i*_ and **y**_*j*_.

The elements of an FRP are determined using three fundamental properties of fuzzy inference:

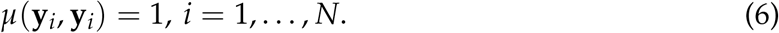

which expresses reflexivity.

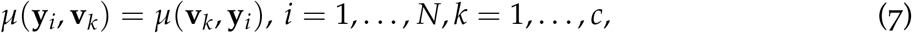

which defines symmetry, where **v**_*k*_ represents the *k*-th cluster center, and *c* > 1 is the specified number of clusters to which each element **y**_*i*_ belongs with an estimated membership level.

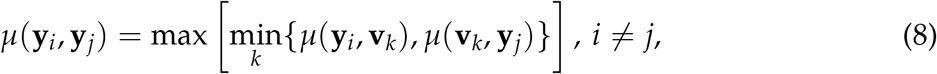

which infers transitivity.

The membership grades *μ*(**y**_*i*_, **v**_*k*_), *i* = 1, …, *N, k* = 1, …, *c*, can be optimally determined using the fuzzy *c*-means (FCM) algorithm [23]. The objective of the FCM algorithm is to minimize the following function:

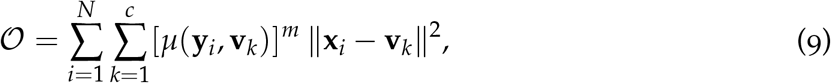

where *m* ∈ [1, ∞), typically set to 2, represents the fuzzy weighting exponent. The FCM objective function is subject to the following constraint:

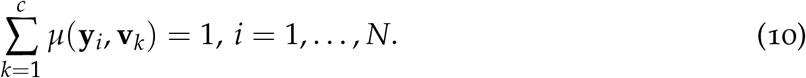

The objective function can be numerically minimized through iterative steps, as follows:

Using initial values for *μ*(**y**_*i*_, **v**_*k*_), *k* = 1, …, *c, i* = 1, …, *N*, the cluster centers and membership grades are iteratively updated until convergence or a predefined number of iterations is reached:

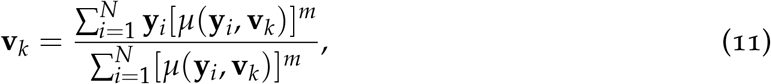

and

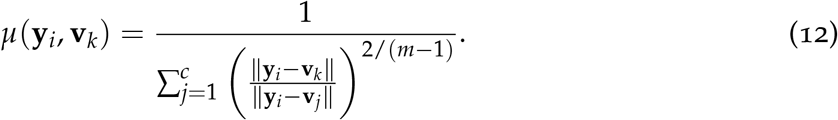

For the computation of the FRPs in this study, the embedding dimension *d*= 1 and time delay *β* = 1 were used for the phase-space construction, and number of clusters *c* = 3 used for the FCM algorithm.

The Shannon entropy of an FRP (FRP-SE), which is a grayscale image, denoted as *H*(**R**), is expressed as

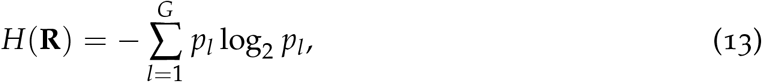

where *G* is set to 256, representing the number of gray levels in **R**. This value is derived by converting real pixel values from the [0, 1] range into integers within [0, 255], *p*_*l*_ corresponds to the probability assigned to the intensity level *l*, which is calculated based on the normalized histogram for the *l*-th bin.

Drawing from the concept of non-probabilistic entropy as defined in [24], the fuzzy recurrence entropy of an *N × N* FRP (FRP-FE), denoted as *Ĥ* (**R**), which quantifies the level of uncertainty in recurrences within the constructed phase space of a dynamical system, is defined as [25]

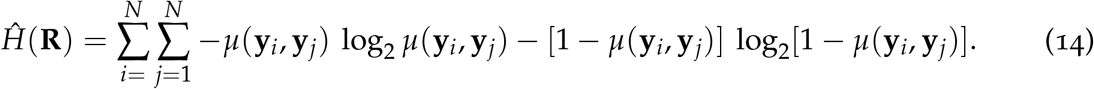

### 3.3 Extraction of wavelet time scattering features

A network for performing wavelet time scattering decomposition employing the analytic Morlet wavelet [26, 27] can be constructed as described in [28]. This network leverages wavelets and a lowpass scaling function to produce low-variance representations of real-valued time series data. The wavelet time scattering process results in representations that remain robust to translations within the input signal while retaining their ability to discriminate between different classes. These representations can be used as inputs to a classifier for pattern classification. The iterative computation of wavelet scattering coefficients across multiple layers is elucidated as follows.

Consider *ψ*(*t*) as a band-pass filter, often referred to as the mother wavelet, which adopts the Morlet wavelet. Additionally, let 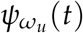 represent a wavelet filter bank, which can be constructed by dilating the mother wavelet as follows:

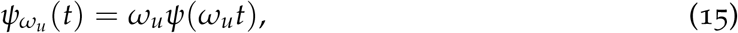

in which *ω*_*u*_ = 2^(*u*/*V*)^, *u* ∈ Z, with 1 ≤ *u* ≤ *U* that is the maximum level of layers, and *V* representing the number of wavelets per octave.

Consider **s** as the input signal. The zeroth-order wavelet scattering coefficients are computed by calculating the average of the feature vector, expressed as

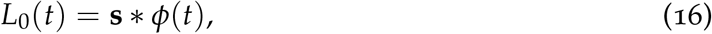

where *L*_0_ represents the zeroth-order scattering, *ϕ* stands for a low-pass filter, and the * symbol denotes the convolution operator.

The coefficients of the first-order wavelet scattering, which belong to layer 1, are calculated by taking the average of the absolute values of the wavelet coefficients at this layer, as follows:

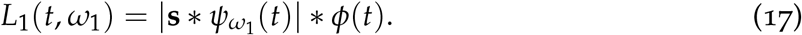

The second-order wavelet scattering coefficients are computed as follows:

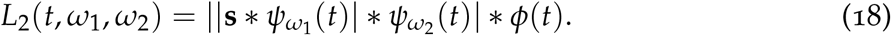

Similarly, the computation of the third-order wavelet scattering coefficients is expressed as

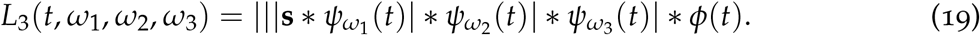

In a general sense, wavelet scattering coefficients at layers *u*, where *u* = 1, …, *U*, are established by applying a combination of convolution, modulus, and average pooling operators as follows:

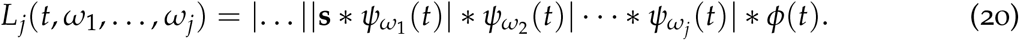

The mother wavelet is defined as [26, 27]

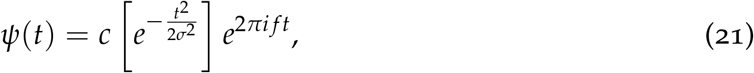

where, in this study, *c* = 1, *σ* is the wavelet duration set to 1, *i* is the imaginary unit, *f* is the central frequency, and 2*π f* is set to 5. Consequently, *e*^2*πi f t*^ = cos(5*t*), resulting in 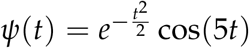.

Other parameters for the wavelet time scattering in this study were specified as follows. Scale of time invariance = half of signal length, using wavelet scattering network with two filter banks, where *V* factor for filter bank 1 = 8 wavelets per octave, and *V* factor for filter bank 2 = 1 wavelet per octave, and sampling frequency = 1 Hz.

### 3.4 Classification with long short-term memory networks

Figure 1 illustrates the progression of an input time series as it passes through an LSTM layer [29]. When utilizing TF and TS features, the input at a given time point is a fusion of four distinct features: IF, SE, FRP-SE, and FRP-FE, all of which are extracted from the corresponding time point segment. Learnable parameters of an LSTM layer encompass the input weights, denoted as **a**, the recurrent weights, denoted as **r**, and the bias, denoted as *b*. These parameters are organized into matrices and vectors as follows: **A** represents the concatenation of input weights, **R** embodies the concatenation of recurrent weights, and vector **b** encapsulates the concatenation of biases from each component. These concatenations are mathematically expressed as:

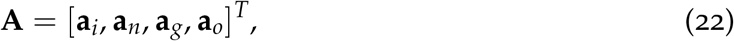

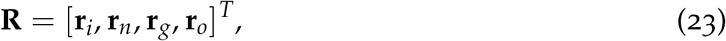

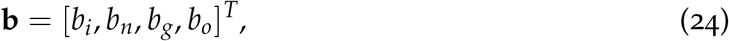

where the subscripts *i, n, g*, and *o* respectively denote the input gate, neglect (or forget) gate, cell candidate, and output gate.

**Figure 1:**
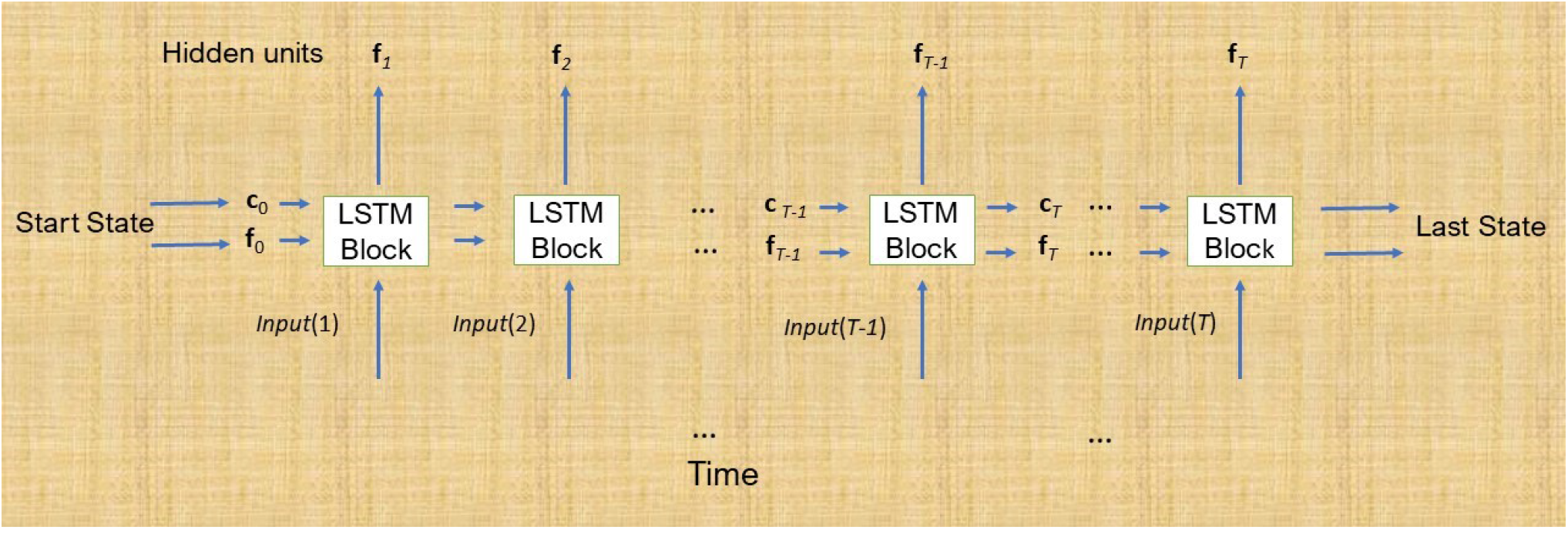
Architecture of a long short term memory (LSTM) network.

The cell state at time step *t* is defined as

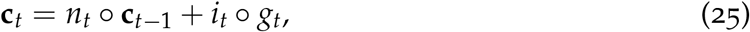

in which ○ is the Hadamard product.

The hidden state at time step *t* is given by

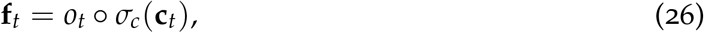

where *σ*_*c*_ represents the state activation function, which is typically computed using the hyperbolic tangent function.

At time step *t*, the input gate (*i*_*t*_), neglect gate (*n*_*t*_), cell candidate (*g*_*t*_), and output gate (*o*_*t*_) are formulated as follows:

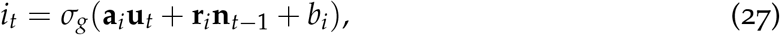

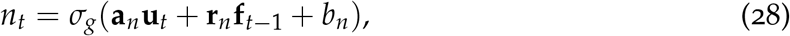

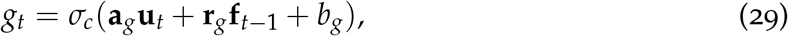

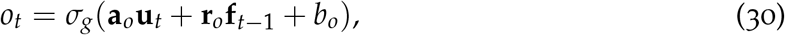

where *σ*_*g*_ represents the gate activation function, commonly employing the sigmoid function.

Moreover, a bidirectional LSTM (bi-LSTM) [30] represents an extension of the LSTM, aiming to offer enhanced performance for sequence classification tasks. Unlike the LSTM, which is trained using a single sequence, the bi-LSTM architecture is trained using two simultaneous LSTM layers: one processes the input time series in its original order, and the other operates on a reversed version of the time series. This dual-layered architecture enables the model to capture bidirectional long-term dependencies between different time steps within the time series data, thereby providing additional contextual information to the network. Consequently, this approach is expected to facilitates more comprehensive learning from the input data.

In this study, the configuration of LSTM and biLSTM networks were chosen as follows. Number of hidden units = 100, output mode = last time step of the sequence, cell and state activation function = hyperbolic tangent, gate activation function = sigmoid. Training options for the networks were set as follow. Solver for training neural network = Adam optimizer, bias learning rate = 1, input-weight *L*_2_ regularizer = nonnegative scalar, recurrent-weight *L*_2_ regularizer = 1, and bias *L*_2_ regularizer = 0.

### 3.5 Measures of classification performance

To compute statistical measures of performance for classifiers, the following variables are defined as

- *P* = the number of samples of pathological voice
- *N* = the number of samples of healthy voice
- *TP* = the numer of correctly predicted samples as pathological voice
- *TN* = the number of correctly predicted samples as healthy voice
- *FP* = the number of falsely predicted samples as pathological voice
- *FN* = the number of falsely predicted samples as healthy voice

The measures of performance in the context of accuracy (*ACC*), sensitivity (*SEN*), specificity (*SPE*), precision (*PRE*), and *F*_1_ score are defined Table 1.

**Table 1:** Statistical performance measures of classification.

| ACC | SEN | SPE | PRE | $F_1$ score |
| --- | --- | --- | --- | --- |
| $\frac{TP + TN}{P + N}$ | $\frac{TP}{P}$ | $\frac{TN}{N}$ | $\frac{TP}{TP + FP}$ | $\frac{2TP}{2TP + FP + FN}$ |

Another performance metric to consider is the area under the receiver operating characteristic (ROC) curve. The ROC curve is generated by plotting the true positive (*TP*) rate against the false positive (*FP*) rate across different thresholds. In some contexts, the TP rate is referred to as sensitivity or the probability of correct prediction, while the FP rate is synonymous with the probability of false alarm. Therefore, the area under the ROC curve (AUC) serves as an assessment of predictor quality, irrespective of the specific classification threshold chosen. Larger values of these performance measures indicate superior performance of the predictor.

## 4 Results

It was shown that the LSTM processing of long signals results in undesired computational complexity in association with the training phase [31]. The original speech signals were divided into short segments of *L* = 250 and 500 time points, where segments whose lengths were less than the specified length were discarded. To overcome the data imbalance, where samples of the pathological conditions are twice more than those of the healthy, partial samples of the healthy speech were replicated to match the sample size of the pathology. Thus, for *L* = 250 and 500, there are 21,746 and 10,861 short speech signals created for each cohort, respectively. Figures 2 and 3 show examples of healthy and pathological speech signals (*L* = 250) and their extracted features, respectively.

**Figure 2:**
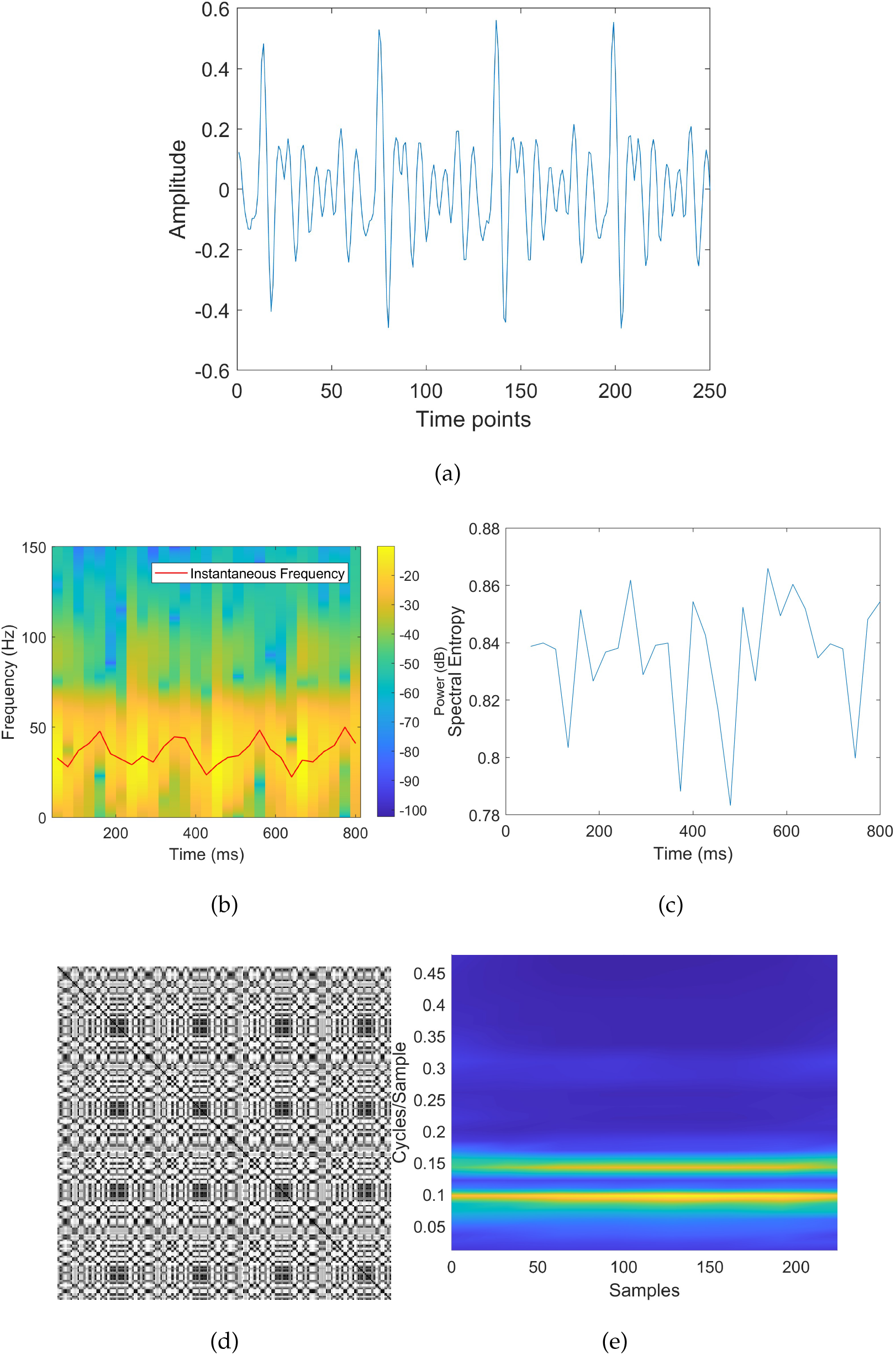
Healthy speech and features: (a) signal segment, (b) instantaneous frequency, (c) spectral entropy, (d) fuzzy recurrence plot, and (e) scattergram of first-order scattering coefficients.

**Figure 3:**
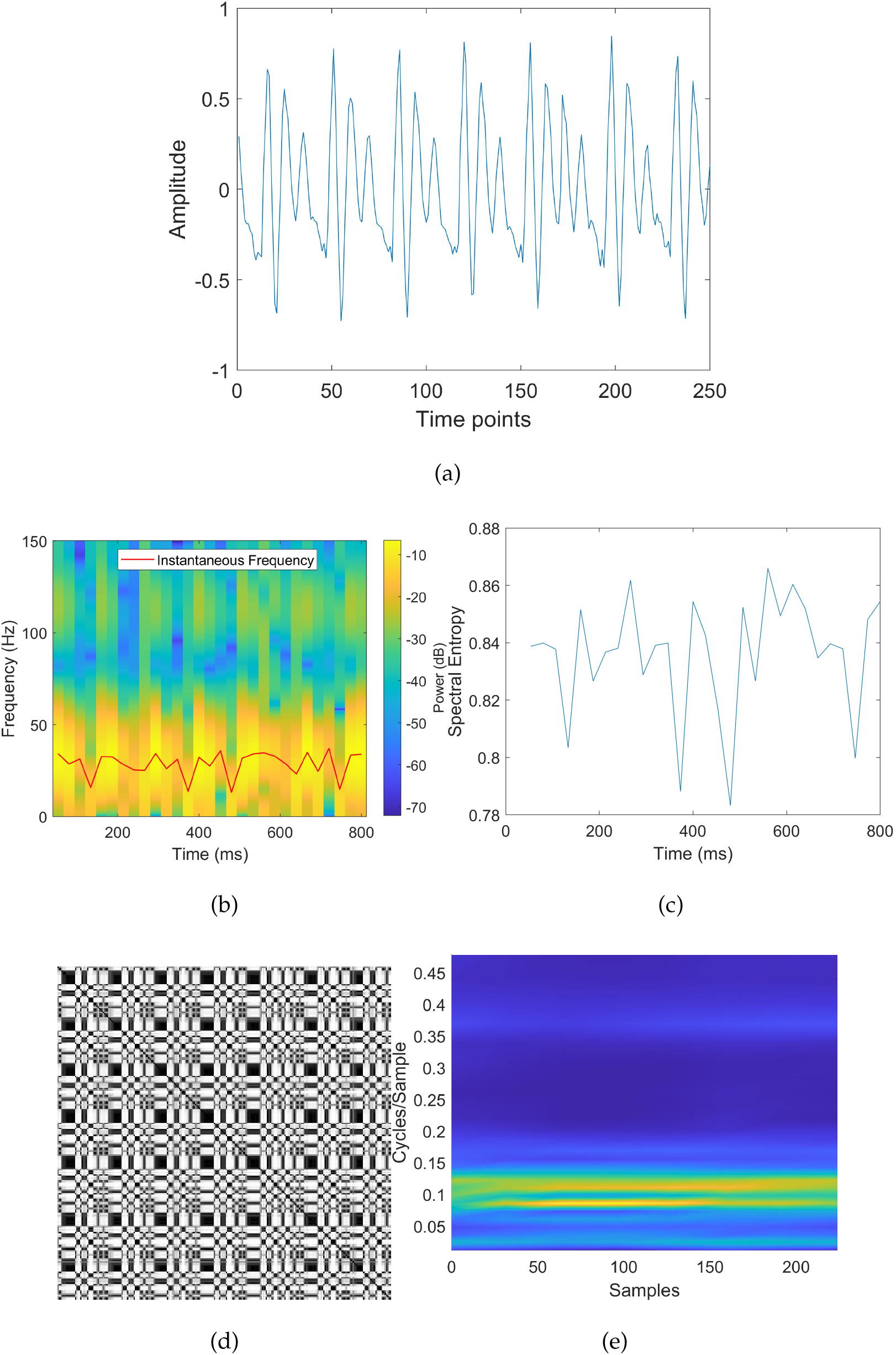
Pathological speech and features: (a) signal segment, (b) instantaneous frequency, (c) spectral entropy, (d) fuzzy recurrence plot, and (e) scattergram of first-order scattering coefficients.

The dataset was randomly split into 10 folds, where 9 folds were used for training the networks and the remaining fold was used for testing the performance of the trained networks. Tables 2 and 3 show results of classifying healthy and pathological voices obtained from different LSTM-based classifiers in terms of accuracy (*ACC*), sensitivity (*SEN*), specificity (*SPE*), precision (*PRE*), *F*_1_ score, and area under the ROC curve (AUC) with respect to *L* = 500 and 250, respectively.

**Table 2:**
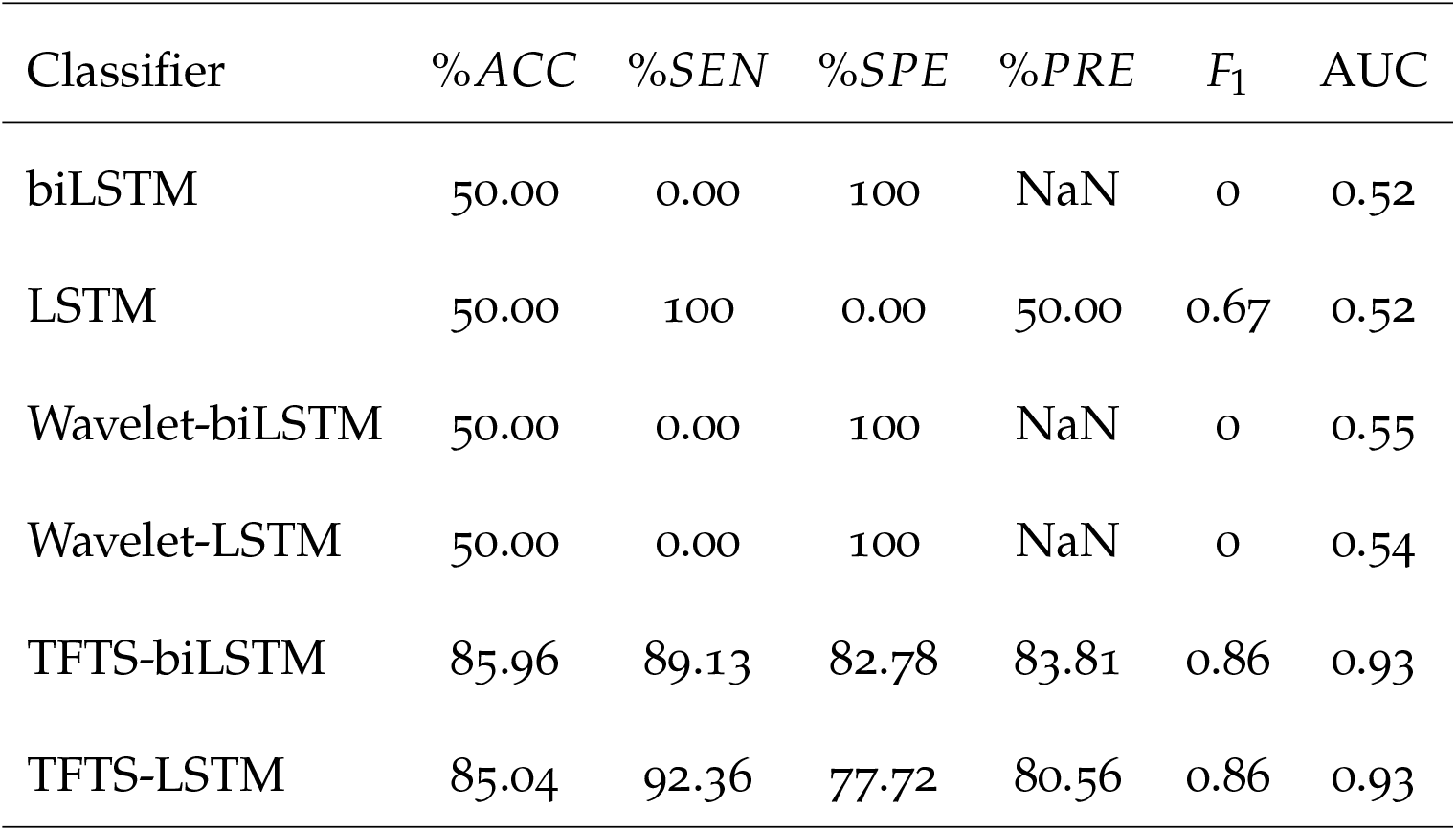
Diagnosis of healthy and pathological speech signals of length = 500 time points.

**Table 3:** Diagnosis of healthy and pathological speech signals of length = 250 time points.

| Classifier | %ACC | %SEN | %SPE | %PRE | $F_1$ | AUC |
| --- | --- | --- | --- | --- | --- | --- |
| biLSTM | 50.00 | 100 | 0.00 | 50.00 | 0.67 | 0.50 |
| LSTM | 50.00 | 0.00 | 100 | NaN | 0 | 0.50 |
| Wavelet-biLSTM | 50.00 | 100 | 0.00 | 50.00 | 0.67 | 0.53 |
| Wavelet-LSTM | 50.00 | 100 | 0.00 | 50.00 | 0.67 | 0.51 |
| TFTS-biLSTM | 88.02 | 93.75 | 82.30 | 84.12 | 0.89 | 0.95 |
| TFTS-LSTM | 90.14 | 93.38 | 86.90 | 87.69 | 0.90 | 0.96 |

For speech segments with *L* = 500, the LSTM, biLSTM, Wavelet-LSTM (LSTM inputs are wavelet time scattering coefficients), and Wavelet-LSTM obtained 50% accuracy and AUC = 0.5. The bi-LSTM, wavelet-LSTM, and wavelet-biLSTM completely biased toward the healthy class (*SPE* = 100%), whereas the LSTM completely biased toward the patholological class (*SEN* = 100%). Both TFTS-LSTM (LSTM inputs are time-frequency and time-space features) and TFTF-biLSTM provided much better results (AUC = 0.93) than the other classification models. Accuracy rates obtained from the TFTS-LSTM and TFTS-biLSTM are 85% and 86%, respectively. Both TFTS-LSTM and TFTS-biLSTM could achieve a balanced classification between the healthy and pathological samples (SEN = 92% and 89%, SPE = 73% and 83%, respectively). The precision measure obtained from the TFTS-biLSTM (84%) is higher than the precision from the TFTS-LSTM (81%), whereas *F*_1_ scores obtained from the two models are the same (0.86).

For shorter speech segments with *L* = 250, complete biases that are opposite to the case with *L* = 500 observed among LSTM, biLSTM, wavelet-LSTM, and wavelet-biLSTM models. The AUC values for LSTM, biLSTM, wavelet-LSTM, and wavelet-biLSTM are between 0.50 and 0.53; whereas the AUC for TFTS-LSTM and TFTS-biLSTM = 0.96 and 0.95, respectively. The use of the shorter signals could improve the performance of both TFTS-LSTM and TFTS-biLSTM models, whose accuracy rates (88% and 90% for TFTS-biLSTM and TFTS-LSTM, respectively) are significantly higher than those of the other four models (50%). Once again, both TFTS-LSTM and TFTS-LSTM could diagnose the pathological conditions better than the healthy samples (*SEN* = 93% vs. *SEN* = 87% for TFTS-LSTM, and *SEN* = 94% vs. *SEN* = 82% for TFTS-biLSTM). The precision and *F*_1_ score of the TFTS-LSTM are higher than those of the TFTS-biLSTM.

Based on the results shown in Tables 2 and 3, the TFTS-LSTM for learning the speech signals of *L* = 250 provided the most favorable classification performance for differentiating between the healthy and pathological voices. To provide a visual comparison between the three different classification methods, Figure 4 shows the iterative machine learning for differentiating the two classes from the speech segments of *L* = 250 and confusion matrices using LSTM, WS-LSTM, and TFTS-LSTM models. It can be seen that both LSTM and WS-LSTM could not improve the training over 50 epochs, whereas the TFTS-LSTM could reach toward nearly the maximum training accuracy or minimum loss.

**Figure 4:**
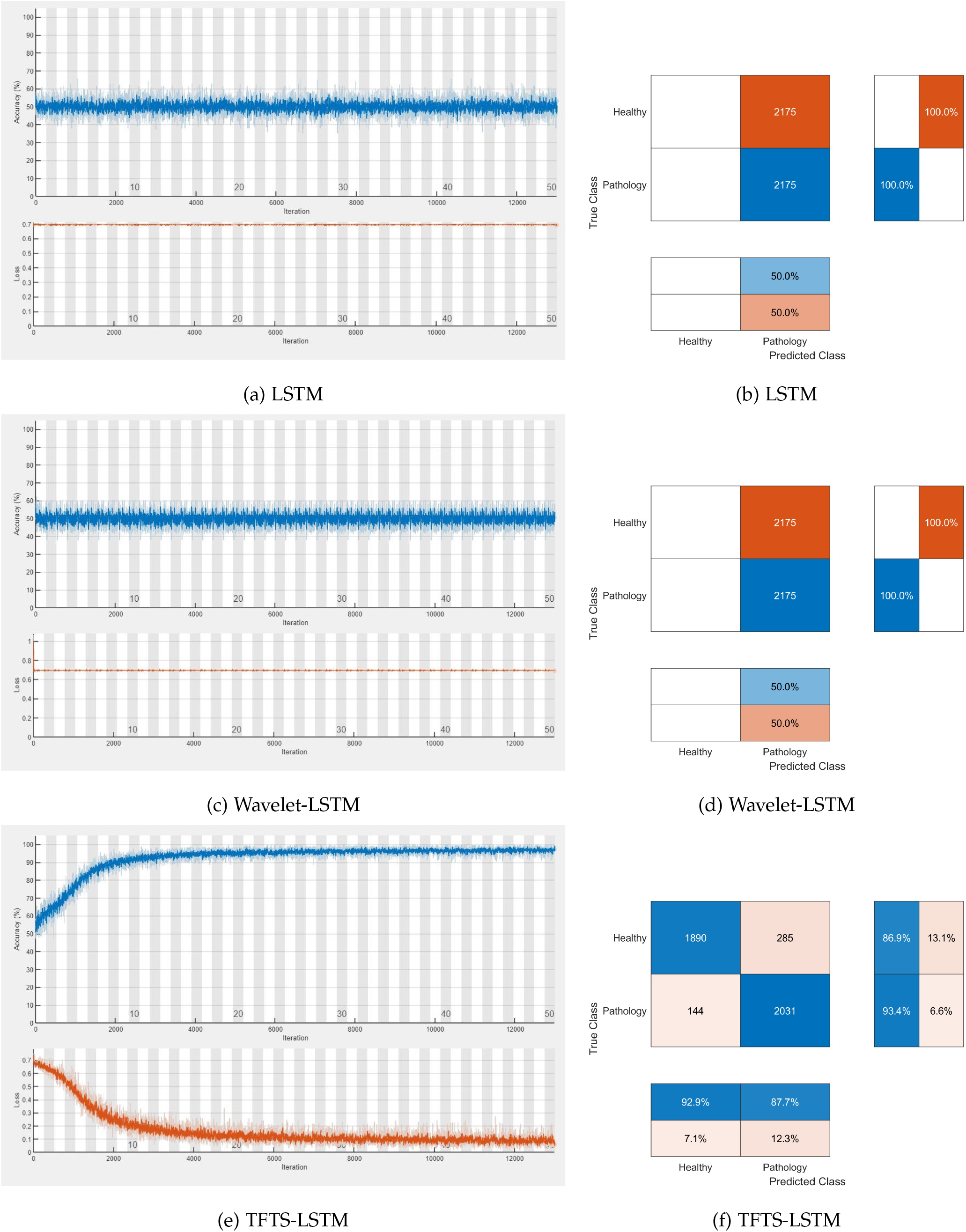
Training processes and confusion matrices using different classification models.

In comparison with using the speech signals of original lengths reported in a previous study [16], in this study, both LSTM and biLSTM increased the classification accuracy to about 2%; wavelet-LSTM and wavelet-biLSTM resulted in about 20% decrease in the classification accuracy; and TFTS-LSTM and TFTS-biLSTM increased the classification accuracy to about 14% and 12%, respectively. A recent study, which incorporated deep-time recurrence features [32] into the LSTM for classifying healthy and pathological speech signals using the same data set, obtained ten-fold results with accuracy = 86%, sensitivity = 100%, and specificity = 50%, showing an unbalanced differentiation between the two cohorts. In these studies, TFTS-LSTM and TFTS-biLSTM outperformed all other methods in terms of accuracy and balance in the diagnosis.

Furthermore, in terms of computational complexity, the use of both LSTM and biL-STM required significant longer training times than the Wavelet-LSTM, Wavelet-biLSTM, TFTS-LSTM, and TFTS-biLSTM. By training these models with a single processor (Intel(R) Core(TM) i7-6500U, <u>CPU@2.50</u> GHz), it was noticed that the time taken for training either the LSTM or biLSTM with the original speech segments were 25 times longer than the time for training the wavelet-LSTM or wavelet-biLSTM, and 10 times longer than the training time for the TFTS-LSTM or TFTS-biLSTM.

## 5 Conclusion

The application of time-frequency and time-space features for LSTM learning was shown to be of high and better performance in classifying physiological signals than several other classification models [19]. In this study, the experimental results illustrated the superior performance of the LSTM that learned on time-frequency and time-space features of healthy and pathological speech signals to those of LSTM networks that learned on either the original data or wavelet-scattering coefficients of the signals.

Certain advantages offered with the incorporation time-frequency and time-space features as input into an LSTM network include better diagnosis accuracy and computational efficiency. A strategy for creating balanced data and augmenting training samples was found to be effective by means of the results obtained from the TFTS-LSTM and TFTS-biLSTM classifiers. In this study, an empirical basis for the selection of short lengths of the speech signals was relied on. An analytical procedure for choosing an optimal signal length is worth developing in a future investigation. Furthermore, considering the inclusion of time-frequency and time-space features in other classifiers would be encouraged for potential improvement of the diagnosis.

Medical voice analysis systems utilize hardware, software, and human-computer interaction to achieve smart hospital facilities [33]. Technical elaborations on this study can contribute to endeavors concerning intelligent technology for the diagnosis of pathology in human acoustics and its potential applications in smart healthcare.

## Data Availability

All data produced are available online at:
https://archive.physionet.org/physiobank/database/voiced/

## Data and Computer Code Availability

The data that were used in this study are publicly available from the PhysioNet website [18]. The Matlab codes implemented in this study are freely available at the first author’s personal website: https://sites.google.com/view/tuan-d-pham/codes, under the title “TFTS-LSTM for pathological voice diagnosis”.

## Declaration of Interest Statement

The authors declare that they have no conflicts of interest.

## Funding

Not applicable.

## Notes

### Competing Interest Statement

The authors have declared no competing interest.

### Funding Statement

This study did not receive any funding

### Author Declarations

VOICED (VOice ICar fEDerico II) database, https://archive.physionet.org/physiobank/database/voiced/

